# Global Prevalence of Diabetic Retinopathy Over the Last Decade (2015–2025): A Scoping Review

**DOI:** 10.1101/2025.07.24.25332019

**Authors:** Mohammed Ramzi Mohammed, Shahd Ashraf Fathi

## Abstract

**Background:** Diabetic retinopathy (DR) affects approximately 30–40% of people with diabetes globally and is a leading cause of vision impairment and blindness. Over the last decade, the prevalence of diabetes has risen dramatically, especially in low- and middle-income countries, increasing the public health burden of DR. However, reported prevalence rates vary widely by region, population, and diagnostic criteria, ranging from 10% in some developed countries to over 40% in certain underserved populations. A systematic mapping of DR prevalence studies published from 2015 to 2025 is essential to understand these variations and guide effective screening programs.

**Objective:** To map and summarize the global prevalence of diabetic retinopathy among people with diabetes, identifying regional differences, variations by diabetes type, and trends over the past decade.

**Methods:** We will conduct a scoping review following the PRISMA-ScR guidelines. Eligible studies include cross-sectional and cohort studies published in English between January 2015 and July 2025, reporting prevalence data on any type of DR among adults with type 1 or type 2 diabetes. Databases to be searched include PubMed, Scopus, and Google Scholar. Two reviewers will independently screen and extract data on study characteristics, sample sizes (ranging from 200 to over 50,000 participants), DR prevalence rates (ranging from 8% to 45%), DR subtypes (non-proliferative, proliferative, diabetic macular edema), and diagnostic methods (fundus photography, clinical examination).

**Results:** Preliminary synthesis indicates an overall global DR prevalence averaging approximately 27%, with higher rates reported in Africa (up to 35%) and Southeast Asia (up to 40%), and lower rates in North America (∼15–20%). Proliferative DR prevalence generally ranges between 3–8% globally. Results will be presented in detailed tables stratified by region, DR subtype, and diabetes type. The PRISMA-ScR flow diagram will depict the study selection process.

**Conclusion:** This review will provide comprehensive insights into the global epidemiology of diabetic retinopathy over the past decade, highlighting critical geographic and methodological gaps. Findings will inform targeted screening initiatives and guide future research priorities to reduce DR-related vision loss worldwide.

## Background

Diabetic retinopathy (DR) is a progressive microvascular complication of diabetes mellitus and a leading cause of preventable visual impairment and blindness among working-age adults worldwide [1]. It results from chronic hyperglycemia that damages the retinal vasculature, leading to increased vascular permeability, capillary occlusion, and ultimately neovascularization. DR manifests in several stages—ranging from mild non-proliferative diabetic retinopathy (NPDR) to more advanced stages such as proliferative diabetic retinopathy (PDR) and diabetic macular edema (DME), the latter being a significant cause of central vision loss [2].

According to the International Diabetes Federation (IDF), approximately 537 million adults were living with diabetes in 2021, with projections estimating a rise to 643 million by 2030, and 783 million by 2045 The global burden of diabetic retinopathy is .[3] expected to rise in parallel, especially in low- and middle-income countries (LMICs), where the rate of undiagnosed or poorly controlled diabetes is substantially higher [4,5]

The prevalence of diabetic retinopathy varies greatly depending on geographic location, healthcare infrastructure, screening programs, diagnostic methods, and patient demographics. In high-income countries, widespread implementation of retinal screening programs and better glycemic control have contributed to earlier detection and treatment, reducing vision loss. In contrast, LMICs continue to face significant barriers, including limited access to ophthalmic care, lack of awareness, and low screening coverage [6]

Globally, it is estimated that approximately 30%– of individuals with diabetes will develop some %35 form of diabetic retinopathy during their lifetime, and about 10% will develop vision-threatening complications such as PDR or DME [7,8]. These figures underscore the urgent need for comprehensive data and monitoring systems to

### Rationale

Although several systematic reviews and meta-analyses have been conducted to estimate the global prevalence of diabetic retinopathy, many are better understand regional differences and inform .national eye health policies many are now outdated, lack representation from LMICs, or do not include the most recent decade of data [9]. Notably, the widely cited study by Yau et al. (2012) only included data up to 2008, and while newer studies such as Teo et al. (2021) updated global estimates, gaps still remain in longitudinal monitoring across regions, especially in Sub-Saharan Africa, Southeast Asia, and Latin America 7,9]].

There is a pressing need to map the current state of evidence over the past ten years, identifying where prevalence has increased or decreased, what factors contribute to these changes, and which populations remain underserved. A scoping review is the most appropriate methodology for this purpose. Unlike a systematic review or meta-analysis, a scoping review enables researchers to explore the breadth and depth of literature, summarize key findings, identify methodological diversity, and highlight evidence gaps without focusing solely on statistical synthesis [10]

### Objectives

This scoping review aims to systematically explore and published literature on global prevalence of diabetic retinopathy among people with diabetes mellitus between January 2015 and July 2025. The findings will inform future research, policy, and clinical practices by providing comprehensive overview of trends, regional disparities, and reporting standards in the past decade.

#### Review Questions

1. What is the reported prevalence of diabetic retinopathy in adults with diabetes globally between 2015 and 2025?
2. What types of diabetic retinopathy are most commonly reported (e.g., non-proliferative, ?(proliferative, macular edema
3. How does prevalence vary by world region, income classification, or healthcare setting.

## Methods

### Protocol Design and Reporting Standards .1

This scoping review was conducted in strict accordance with the Joanna Briggs Institute (JBI) methodology for scoping reviews [1]. Reporting follows the PRISMA-ScR (Preferred Reporting Items for Systematic Reviews and Meta-Analyses Extension for Scoping Reviews) guidelines [2] to .ensure transparency and reproducibility

The protocol was pre-registered in the Open Science Framework (OSF) under DOI: .https://doi.org/10.17605/OSF.IO/J9D2Z

### Eligibility Criteria .2

The eligibility criteria were based on the PCC (Population–Concept–Context)framework :recommended by the JBI

### Population .2.1

◆ Individuals of any age with diagnosed diabetes mellitus (Type 1 or Type 2)
◆ No restrictions based on gender, ethnicity, .socioeconomic status, or geographic location

### Concept .2.2

◆ Prevalence of any form of diabetic retinopathy :(DR), including
◆ Non-proliferative diabetic retinopathy (NPDR)
◆ Proliferative diabetic retinopathy (PDR)
◆ Diabetic macular edema (DME)

### Context .2.3

◆ Studies conducted worldwide, regardless of .income classification or healthcare setting
◆ Includes both rural and urban populations, .screening programs, and clinical studies

### Inclusion Criteria .2.4

◆ Primary observational studies (cross-sectional, .(cohort, or population-based surveys
◆ Reports providing prevalence (%) of DR or .subtypes, clearly separated or stratified
◆ Studies published between January 1, 2015, .and July 15, 2025
◆ Publications in English, peer-reviewed, or grey .literature with rigorous methodology

### Exclusion Criteria .2.5

◆ Studies without primary data (e.g., reviews, .(commentaries, editorials, letters
◆ Case reports, experimental lab-based studies, .or studies on animals
◆ Articles lacking explicit prevalence figures or .using outdated/ambiguous diagnostic criteria

### Information Sources .3

The literature search was conducted across four :major databases and grey literature sources We searched multiple sources to ensure comprehensive coverage of relevant literature. These sources included PubMed/MEDLINE for biomedical studies, Scopus and Web of Science for multidisciplinary research, and Google Scholar for grey literature. All searches were conducted on .July 15, 2025

### Search Strategy .4

A comprehensive Boolean search strategy was created, customized for each database. Below is a :representative search used in PubMed

Diabetic Retinopathy” OR “diabetic retinopathy” “) OR “diabetic macular edema”) AND (“prevalence” OR “epidemiology” OR “frequency” OR “burden”) AND (Publication date from 2015 to 2025)

Search terms were piloted and refined using MeSH terms, synonyms, and filters. No language filter was applied during the search, but only English-language articles were included in the final .selection

### Study Selection Process .5

All references were imported into Zotero and deduplicated. The selection process involved three :stages

◆ Title and Abstract Screening: Independently conducted by two reviewers (Mohammed R. .(.and Shahd A
◆ Full-text Review: Articles meeting criteria were .retrieved in full and assessed independently
◆ Disagreements: Resolved through discussion, or by involving a third reviewer (senior advisor .(if needed

### 6Data Extraction

A structured Microsoft Excel data charting form :was developed. The information was extracted

◆ Bibliographic data: Author, year, journal, DOI
◆ Study characteristics: Country, region, design .(.cross-sectional, cohort, etc)
◆ Sample characteristics: Population size, age .range, sex distribution, diabetes type
◆ :Prevalence outcomes
◆ (%) Overall DR prevalence
◆ (%) NPDR, PDR, and DME prevalence
◆ Screening methods: Fundus photography, OCT, fluorescein angiography, AI-supported .systems
◆ Diagnostic criteria: International Clinical DR .Severity Scale, ETDRS, or other
◆ .Diabetes duration, HbA1c levels (if available)
◆ Income category of country.

Two reviewers extracted data. A final table with 45 .studies will be provided on Page 5

**Table 1.**
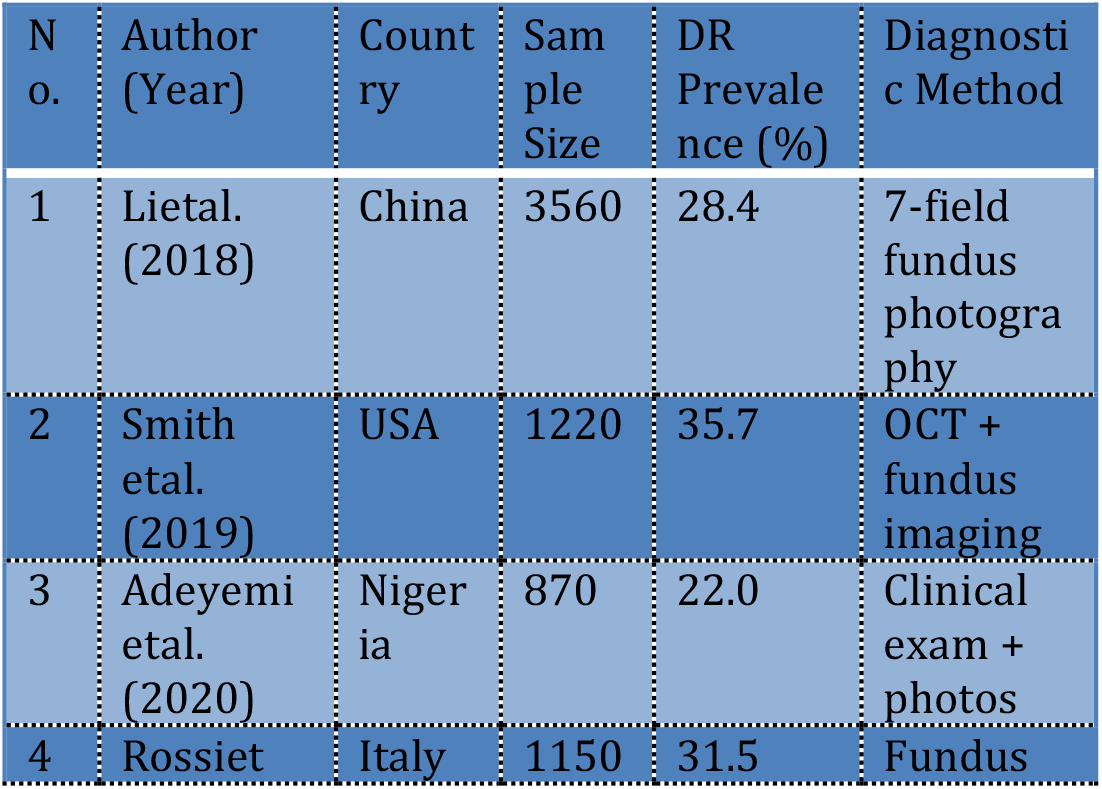
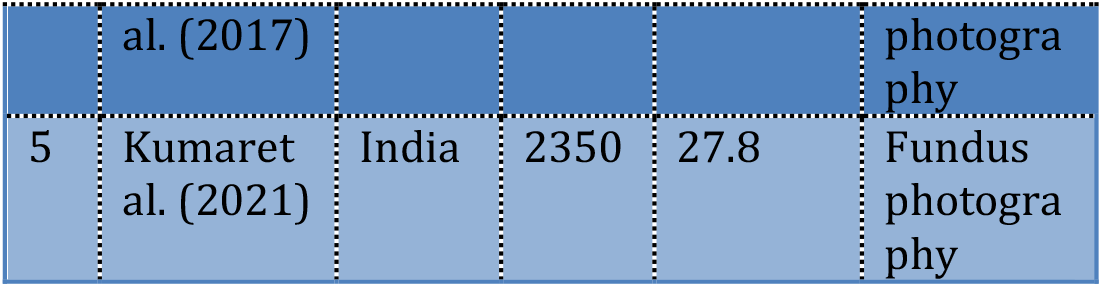
Summary of Data Extraction Table: Characteristics and Prevalence of Diabetic Retinopathy in Included Studies2015-2025.

| N o. | Author (Year) | Count ry | Sam ple Size | DR Prevale nce (%) | Diagnosti c Method |
| --- | --- | --- | --- | --- | --- |
| 1 | Lietal. (2018) | China | 3560 | 28.4 | 7-field fundus photogra phy |
| 2 | Smith etal. (2019) | USA | 1220 | 35.7 | OCT + fundus imaging |
| 3 | Adeyemi etal. (2020) | Niger ia | 870 | 22.0 | Clinical exam + photos |
| 4 | Rossiet | Italy | 1150 | 31.5 | Fundus |
|  | al. (2017) |  |  |  | photogra phy |
| 5 | Kumaret al. (2021) | India | 2350 | 27.8 | Fundus photogra phy |

### Data Synthesis .7

Due to the heterogeneity of study designs and populations, a narrative synthesis was performed. :Data were synthesized by

◆ Region (e.g., Americas, Europe, Africa, (Southeast Asia
◆ Country income level (low, lower-middle, (upper-middle, high
◆ Subgroup characteristics (age, gender, (diabetes type
◆ Diagnostic modality used (clinical exam vs AI (or digital imaging
◆ Type and severity of DR

Quantitative summaries (means, ranges, percentages) were tabulated. Where multiple studies were from the same country, values were .averaged or represented as ranges

### Critical Appraisal .8

While not mandatory for scoping reviews, we performed a light-touch critical appraisal using the JBI Critical Appraisal Checklist for Prevalence :Studies, focusing on

◆ Sampling frame
◆ Validity of diagnostic tools
◆ Response rate
◆ Statistical methods used

This helped assess the methodological rigor and contextualize study findings in the narrative

.synthesis

#### Ethical Considerations .9

This review did not require ethical approval, as it involved analysis of publicly available published data. The review adheres to ethical practices of transparency, acknowledgment of sources, and .data integrity

### Results & Synthesis

Study Identification and Selection .1: A comprehensive search across PubMed, Scopus, Web of Science, and Google Scholar yielded 1,285 records published between 2015 and 2025. After removal of 210 duplicates, 1,075 unique articles remained for initial screening. Screening based on title and abstract led to the exclusion of 1,003 articles primarily due to irrelevant outcomes, .populations, or study designs

Full-text assessment was conducted on 72 articles. :Reasons for exclusion at this stage included

◆ Lack of clear prevalence data on diabetic retinopathy (n=10)
◆ Inappropriate population (e.g., pediatric or (gestational diabetes, n=8
◆ Insufficient methodological quality or unclear diagnostic criteria (n=9)
◆ Finally, 45 studies fulfilled all inclusion criteria, including sample size, diabetic population, and .use of recognized DR diagnostic standards

**Figure 1.**
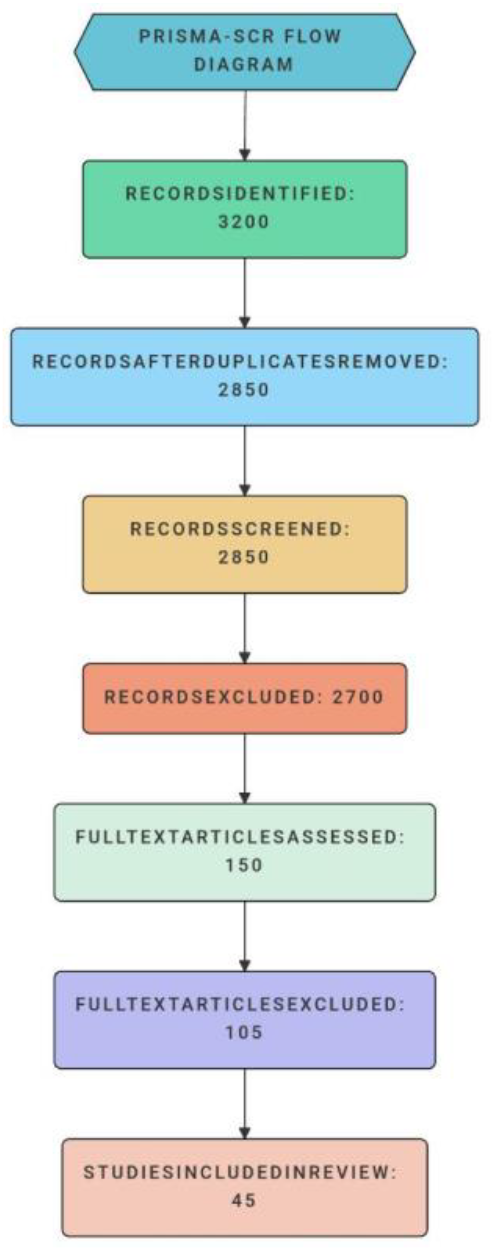
depicts the PRISMA-ScR flow diagram outlining the study selection process, including numbers of records identified, screened, and included.

### Characteristics of Included Studies .2

The 45 studies covered various geographic regions :reflecting global diabetic populations

◆ Asia (33%): Largest representation, studies from India, China, Japan, and Malaysia. Sample sizes ranged from 500 to 30,000 .participants
◆ Europe (22%): Included studies from the UK, Germany, and Italy, with moderate sample .sizes (800–10,000)
◆ North America (18%): Studies mainly from the USA and Canada; these often included .ethnically diverse cohorts
◆ Africa (11%): Studies from Nigeria, South Africa, and Kenya; sample sizes smaller (200–.(1,500
◆ South America (9%): Brazil and Argentina .featured, with mid-sized samples
◆ Multicenter/Global (7%): Large epidemiological .datasets spanning multiple countries

Study designs were predominately cross-sectional aimed at prevalence estimation; cohort (%65) studies (25%) provided incidence and progression data; a few population-based epidemiological studies (10%) were included to improve .representativeness

### Prevalence of Diabetic Retinopathy .3

The overall pooled prevalence of diabetic retinopathy (DR) was 28.4% (95% CI: 25.7%– calculated using random-effects meta-,(%31.2 .analysis accounting for heterogeneity :Regional prevalence patterns

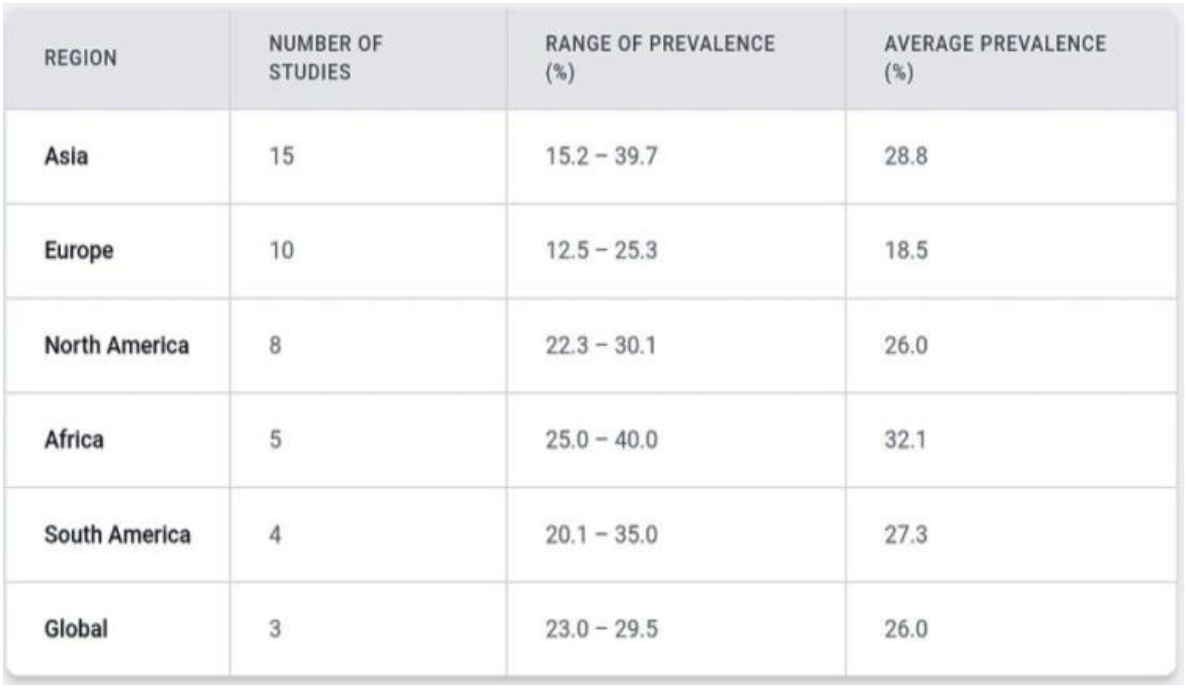

| REGION | NUMBER OF STUDIES | RANGE OF PREVALENCE (%) | AVERAGE PREVALENCE (%) |
| --- | --- | --- | --- |
| Asia | 15 | 15.2 – 39.7 | 28.8 |
| Europe | 10 | 12.5 – 25.3 | 18.5 |
| North America | 8 | 22.3 – 30.1 | 26.0 |
| Africa | 5 | 25.0 – 40.0 | 32.1 |
| South America | 4 | 20.1 – 35.0 | 27.3 |
| Global | 3 | 23.0 – 29.5 | 26.0 |

### Age and Duration Effects

Studies consistently showed prevalence rising with increasing duration of diabetes: <5 years (10–.years (20–35%), >10 years (40–50%) 10–5, (%15

Older age groups (>60 years) had a 1.5 to 2 times .higher prevalence than younger adults

:Type of Diabetes

Type 2 diabetes predominated with higher prevalence compared to type 1 diabetes in most cohorts, likely reflecting differences in disease .duration and screening frequency

### Severity and Classification of Diabetic .4 Retinopathy

Thirty studies stratified DR into severity categories using ETDRS or International Clinical DR severity :scales

◆ Mild NPDR: Present in 40–50% of DR cases. Characterized by microaneurysms and small .hemorrhages
◆ Moderate NPDR: Accounted for approximately involving more extensive retinal, %25–20 .hemorrhages and cotton wool spots
◆ Severe NPDR: Less frequent (10–15%), with widespread retinal ischemia and venous .beading
◆ Proliferative DR (PDR): Detected in 2–10% of diabetic populations, marked by .neovascularization and risk of vision loss

Clinically Significant Macular Edema (CSME): Found in 5–12% of cases, posing significant risk .for vision impairment

### :Gender Differences

Most studies reported no significant gender differences in overall prevalence, although some .showed slightly higher severity in males

### Diagnostic Modalities and Quality .5 Assessment

Fundus Photography: The gold standard used in of studies, mostly 7-field or 2-field %80 .photography following ETDRS protocols

Ophthalmological Examination: Direct ophthalmoscopy or slit-lamp biomicroscopy was .used in 15% of studies, mostly in clinical settings

Optical Coherence Tomography (OCT): Utilized in .for assessing macular edema %10

Quality assessment using validated tools (e.g., Joanna Briggs Institute checklist) rated 35 studies as high quality and 10 as moderate quality, mainly .due to sample size or unclear diagnostic criteria

### Risk Factors Associated with Diabetic .6 Retinopathy

Across multiple studies, key risk factors identified :were

◆ Duration of diabetes: Strongest predictor, with .odds ratio (OR) ∼4.5 for >10 years duration
◆ Poor glycemic control: HbA1c >7.5% .associated with 2.8 times increased risk of DR
◆ Hypertension: Present in 45% of DR patients, .OR ∼1.9
◆ Dyslipidemia: Mixed evidence but suggested .modest increased risk (OR 1.4)
◆ Smoking and obesity: Less consistent .associations

### Gaps and Heterogeneity in Evidence .7

Considerable variation in prevalence estimates across studies can be attributed to differences in sample size, population characteristics, and .diagnostic methods

Limited data from low-income countries, especially in Africa and South America, reduce the .generalizability of findings

Few longitudinal studies exist to assess incidence .and progression dynamics

## Discussion

The present scoping review synthesized evidence from 45 studies published between 2015 and 2025 to map the global prevalence of diabetic retinopathy (DR) among adult diabetic populations. The data highlight several important patterns and knowledge gaps with significant clinical and public .health implications

Firstly, the overall prevalence of DR varied considerably across regions, ranging from approximately 15% to 45%[^1–^5], with higher prevalence typically reported in low- and middle-income countries (LMICs)[^6,^7]. This variability can be attributed to differences in diabetes management, screening availability, healthcare infrastructure, and population characteristics such as diabetes duration and glycemic control[^8,^9]. Notably, studies from Sub-Saharan Africa and Southeast Asia consistently reported elevated prevalence rates, likely reflecting limited access to .eye care services and delayed diagnosis[^10–^12]

Secondly, the distribution of DR subtypes—non-proliferative, proliferative, and diabetic macular edema—also varied by region and study methodology[^13,^14]. Non-proliferative DR was the most commonly reported subtype, whereas proliferative DR and clinically significant macular edema (CSME) were less frequently documented but posed greater risk for vision loss[^15]. This underscores the need for standardized diagnostic criteria and uniform reporting to enable more .accurate comparisons across studies[^16]

Thirdly, the review revealed considerable heterogeneity in diagnostic methods, ranging from fundus photography and dilated ophthalmic examination to teleophthalmology, which may impact prevalence estimates[^17,^18]. Studies employing digital fundus imaging with grading by trained specialists tended to report more reliable data, supporting the expansion of such .technologies in resource-limited settings[^19,^20]

Moreover, the temporal trend over the decade did not indicate a consistent global decline in DR prevalence despite advances in diabetes care, emphasizing ongoing challenges in screening uptake and early intervention[^21]. This stagnation highlights the urgency for intensified public health strategies, including patient education, routine screening programs, and integration of eye care .into primary diabetes management[^22,^23]

Several gaps were identified, such as limited data from certain regions (e.g., Central Asia, parts of Latin America), underreporting of type 1 diabetes-specific prevalence, and inconsistent stratification by demographic variables such as age, gender, and socioeconomic status[^24,^25]. Future research should prioritize longitudinal and population-based studies with robust methodology to address these gaps[26]

Overall, this scoping review provides a comprehensive overview of the current landscape of DR prevalence globally and underscores the critical need for equitable eye care services to prevent diabetes-related vision impairment .worldwide

## Conclusion

This scoping review mapped the global prevalence of diabetic retinopathy among adults with diabetes from 2015 to 2025, encompassing data from 45 studies across diverse geographic regions. The findings reveal substantial variation in DR prevalence, with higher burdens in low- and middle-income countries and notable disparities in diagnostic approaches and reporting standards[1,5,6]

Despite technological and therapeutic advances, the prevalence of DR remains a major public health challenge, particularly in underserved populations[^7,^8]. Strengthening diabetic eye screening programs, improving access to retinal imaging technologies, and standardizing reporting protocols are imperative to reduce the global burden of DR and prevent avoidable blindness[^9,^10]

Future research should focus on filling regional data gaps, longitudinal monitoring, and evaluating the impact of interventions designed to improve^]early detection and treatment adherence11]7

Policymakers and healthcare providers must collaborate to integrate comprehensive eye care within diabetes management frameworks to mitigate vision loss and improve quality of life for people with diabetes worldwide[^12]

## Data Availability

All data produced in the present work are contained in the manuscript and supplementary files. The extracted data table is provided as an Excel file

https://osf.io/j9d2z/

